# Validating data from Multiplex Assays of Variant Effect (MAVEs): A CanVIG-UK National Survey of NHS Clinical Scientists

**DOI:** 10.1101/2025.02.07.25321851

**Authors:** Sophie Allen, Alice Garrett, Charlie F. Rowlands, Miranda Durkie, George J. Burghel, Rachel Robinson, Alison Callaway, Joanne Field, Bethan Frugtniet, Sheila Palmer-Smith, Jonathan Grant, Judith Pagan, Trudi McDevitt, Katie Snape, Helen Hanson, Terri McVeigh, David J. Adams, Gregory M. Findlay, Rehan M. Villani, Amanda B. Spurdle, Clare Turnbull, CanVIG-UK

## Abstract

Advances in technology have made possible Multiplex Assays of Variant Effect (MAVEs), systematically generating functional data for many thousands of genetic variants. Robust clinical validation and accessible online resources for MAVE data have previously been identified as barriers to clinical adoption of new MAVEs.

We delivered a survey during the November 2024 Cancer Variant Interpretation Group (CanVIG-UK) meeting comprising NHS clinical scientists and clinical geneticists, and received 46 responses from individuals regularly performing variant classification for diagnostic reporting.

Only 35% reported they would accept clinical validation of the MAVE provided by its authors: 20% reported they would attempt clinical validation themselves whilst 61% would await clinical validation by a trusted central body. 72% reported they would use MAVE data ahead of formal peer-reviewed publication if reviewed and clinically validated by a trusted central body. When scoring such central bodies on a scale 1-5 for confidence in their review and validation of MAVEs, CanVIG-UK (median=5), VCEPs (median=5) and ClinGen SVI Functional Working Group (median=4) all scored highly. Participants supported making variant-level data accessible via a relevant web resource (although the majority of participants expressed that additional assay-level or variant-level information would have a low likelihood to alter validation scores provided by a trusted central body). These findings, from a comparatively homogenous clinical diagnostic group operating in a resource-constrained healthcare setting, indicate that clinical application of new MAVEs for variant classification will be delayed unless robust clinical validations are performed by a trusted central body and made readily accessible.

## Introduction

Rapid advances in high-throughput molecular and bioinformatic pipelines for genomic sequencing have shifted the bottleneck in diagnostic genetic testing to the classification and clinical interpretation of detected variants. On encountering a new, rare variant on clinical testing, population-level, case-control, segregation and phenotype-related data from patients with the disease are frequently insufficient to enable robust classification as pathogenic or benign. Thus, this may result in classification as a variant of uncertain significant (VUS), a problem exacerbated in patients of non-European descent on account of smaller available population-level datasets^1–3^. Assays of variant impact on gene function provide an attractive alternative information source for differentiating which variants are deleterious and which are neutral with respect to function. Historically, functional assays were low-throughput, typically only reporting on a handful of repeatedly-observed variants. Through concurrent advances in high throughput sequencing and gene-editing technologies, so-called Multiplexed Assays of Variant Effect (MAVEs) can provide systematically-generated data for many thousands of variants, enabling ‘pre-annotation’ of (nearly) all variants that might ever be observed clinically^4,5^.

Robust validation of the assay across the variant types deemed causative to the corresponding phenotype is critical to clinical application for variant interpretation of data from MAVEs, or any functional assays. For genes where disease is conferred through loss-of-function, this usually comprises analysing the separation of assay readouts between synonymous and nonsense variants: this both validates performance of the assay in predicting loss-of-function and allows calibration of thresholds for designation of impact (or not) on function. However, under the 2015 American College of Medical Genetics and Genomics (ACMG) and Association for Molecular Pathology (AMP) variant classification framework, truncating and synonymous variants are less likely to be reliant on novel functional data to attain definitive classifications of pathogenic or benign^6^. Rare missense variants (which are in aggregate quite frequent) are more commonly problematic for confident classification as pathogenic or benign. Hence, for these variants it is important that the assay has been validated for its ability to distinguish between known benign and pathogenic missense variants. Methodology quantifying concordance of assay calls against truthsets of benign and pathogenic variants for the relevant ACMG/AMP codes for pathogenicity (PS3) and benignity (BS3) was approved by the ClinGen Sequence Variant Interpretation (SVI) Functional Working Group (WG) and published in 2019 by Brnich et al^7^.

We previously conducted a workshop in July 2023 at the Wellcome Trust Sanger Institute in Cambridge, UK, bringing together invited international stakeholders from the ClinGen community (attending the 2023 Curating the Clinical Genome meeting) and from the Atlas of Variant Effects (AVE) Alliance community (attending the Mutational Scanning Symposium)^8^. In this half-day workshop, we sought to explore barriers and facilitators to clinical adoption of MAVEs. Two predominant issues emerged: (i) standards for Brnich-style validation of MAVEs, and organisational approaches by which these validations might be undertaken in a timely, accurate, consistent and efficient fashion, and (ii) standards for release of and platforms for clinical display of MAVE data.

A number of international bodies were recognised as potentially relevant to setting standards and/or delivery of Brnich-style validation of MAVEs. The ClinGen SVI Functional WG notably established the initial Brnich et al. methodology (https://www.clinicalgenome.org/working-groups/sequence-variant-interpretation). The Atlas of Variant Effects (AVE) Alliance was established in 2020 by the community working in MAVEs and has a Clinical Variant Interpretation (CVI) workstream, again established broadly for the same goal^9^. Variant Curation Expert Panels (VCEPs) have been established by ClinGen: these voluntary, expert-led groups develop (typically over 1-4 years) individual gene-level specifications of the 2015 ACMG/AMP variant classification framework^10^.

The major options for deposition of MAVE data (beyond individual publications and archived pre-prints) were recognised to be (i) ClinVar and (ii) MaveDB^11,12^. There have been comparatively low rates of deposition of variant-level assay/MAVE data into ClinVar to date, but as a platform ClinVar has the advantage of being very familiar to the clinical diagnostic community. MaveDB was established by the AVE Alliance as a repository of MAVE data targeted primarily towards the MAVE-generating scientific community (and is not currently searchable on a per-variant basis).

In the UK, following adoption of the 2015 ACMG/AMP variant classification framework and at the request of the British Association of Genomic Medicine (BSGM) and Association of Clinical Genomic Science (ACGS), the Cancer Variant interpretation Group-UK (CanVIG-UK) was established in 2017, with monthly meetings held since that date^13^. Membership comprises 411 National Health Service (NHS) Clinical Scientists, Consultants, Clinical Geneticists, and Genetic Counsellors from the UK and Republic of Ireland working in the field of cancer susceptibility genetics. An oversight group, the CanVIG-UK Steering Advisory Group (CStAG), also meets monthly, comprising 13 lead clinicians and clinical scientists providing representation across each regional laboratory group (Genomic Laboratory Hub (GLH)) alongside the central CanVIG-UK team (2-3 clinical-research trainees, variously supported by research funding). The goal of CanVIG-UK and CStAG is to generate resources to support consistency in the UK for ACMG/AMP-based variant classification for cancer susceptibility genes, with activities including educational meetings, consultative forums for problematic variants, development of resources such as templates for clinical diagnostic reporting and development/specification/ratification of guidance for variant interpretation. One resource developed by CanVIG-UK has been a centralised database capturing Brnich-style assay validations performed by the CanVIG-UK central team and collectively via the CanVIG-UK clinical scientist community (https://www.cangene-canvaruk.org/functional-studies-recommendations). Another resource developed and maintained by the CanVIG-UK central team is CanVar-UK, an online platform established in 2017 that hosts variant-level dialogue classifications from the CanVIG-UK clinical scientist community as well as multiple annotations for >1,100,000 variants in 116 cancer susceptibility genes, including data from 23 relevant MAVEs/functional assays (https://canvaruk.org/).

Recognising the challenges relating to clinical validation and data access for MAVEs, alongside the rapid increase in publication of these assays, we sought to capture perspectives on these issues directly from those performing ACMG/AMP-based variant classification in a ‘real-world’, financially constrained healthcare setting. We therefore undertook a structured survey of the NHS clinical scientists and clinical geneticists through CanVIG-UK.

## Subjects and Methods

Question content and electronic format/usability was iterated through several expert groups knowledgeable in ACMG/AMP-based variant classification, to ensure questions were clear for participants with a clinical background. These groups included The CanVIG-UK Steering and Advisory Group (CStAG; MD, GJB, RR, AC, JF, BF, SPS, JG, JP, TMcD, KS, HH, and TMcV) and members of the Atlas of Variant Effect Alliance Clinical Variant Interpretation Workstream (AVE-CVI; AS and RV) (https://www.varianteffect.org/workstreams) (Supplementary Methods).

Participants were attendees at the CanVIG-UK monthly meeting on 15^th^ November 2024 who self-selected to participate. Survey data was collected online (http://www.surveymonkey.com), the link to which was circulated to attendees both during the meeting and afterwards by email and remained live until 20^th^ November 2024. To provide all participants with an equivalent base-level of knowledge regarding MAVE data, the meeting consisted of four talks to introduce the topics surveyed, comprising (i) validation of MAVE data and the Brnich et al. methodology^7^, (ii) resources and repositories for accessing MAVE data, including MaveDB, ClinVar and CanVar-UK, and (iii) talks from authors (GF, DA) on recently published MAVEs for *VHL* and *BAP1*^14,15^.

The survey comprised 11 questions relating to use of MAVE data in NHS clinical diagnostic labs and covered (i) performing clinical validation of MAVE data locally within their own laboratory, (ii) processes for clinical validation of MAVE data: centralised versus local, and (iii) gene-level and variant level-data required additional to PS3/BS3 assay-level scores for clinical application of MAVE data (Table S1). The survey received a total of 48 responses of the 86 attendees present at the CanVIG-UK meeting (recruitment rate: 55.8%), all of which were from unique IP addresses. 46/48 responses were from individuals who confirmed regular undertaking of ACMG/AMP-based variant classification for clinically diagnostic reporting, and these 46 results were retained for further analysis.

Figures, counts, and summary statistics were generated using R (version 4.3.0 (2023-04-21 ucrt)^16^) and R Studio (version 2024.12.0+467^17^).

## Results

### Participants

Of the 46 participants who confirmed they regularly perform ACMG/AMP-based variant classification for clinical diagnostic reporting, the majority (43/46) were clinical scientists and the remainder (3/46) were clinical genetics consultants.

### Performing clinical validation of MAVE data locally within own laboratory

When asked if they had ever undertaken a Brnich-style validation of any functional assay, just under half of participants (20/46, 43.5%) responded ‘Yes’ (Figure 1A). Participants were then asked, should a new MAVE be published that they would deem to be useful for variant classifications for a gene that they report in their local GLH, whether they would attempt a Brnich-style validation of the MAVE. Only 9 (19.6%) reported that they would attempt the Brnich-style validation of a MAVE themselves. Of the other 37 respondents, 2 (4.3%) stated that they would anticipate that another scientist in their GLH would undertake the validation, 28 (60.9%) would await validation by a central body such as CanVIG-UK or the VCEP, and 7 (15.2%) were unsure of their local processes for validation of functional assays (Figure 1B).

**Figure 1:**
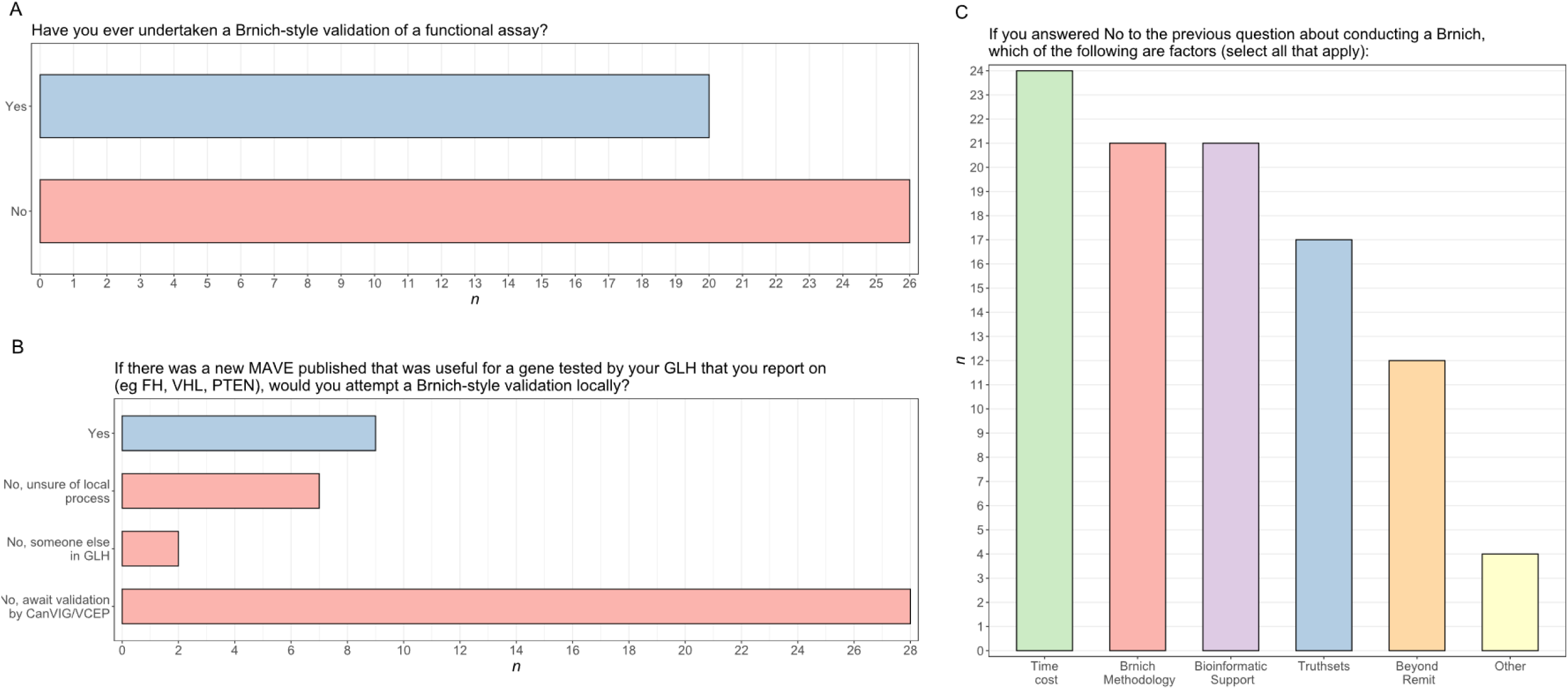
Results from survey questions pertaining to local clinical validation. *n* = Number of participants. A: Number of participants reporting if they have or have not ever attempted a Brnich-style validation themselves (n=46). B: Number of participants who would or would not undertake Brnich-style validation for a new MAVE of utility for a gene (n=46). C: Reasons why participants would not attempt a Brnich-style validation locally (n=37). Participants could select one or more of the given options for this question.

Reasons cited by the 37 respondents who would not attempt a Brnich-style validation themselves included the process being too time-consuming (24/37), their lack of confidence in the validation methodology (21/37), lack of confidence in defining the truth-sets (17/37) and lack of available bioinformatic support within their GLH (e.g. for extracting truthset variants from ClinVar and aligning with assay results, 21/37). 12/37 responded that they considered Brnich-style validation beyond the remit of their role (Figure 1C).

### Processes for clinical validation of MAVE data: centralised versus local

Next, participants were asked to consider the situation of authors of a new MAVE having included a Brnich-style validation with assay-level PS3/BS3 scores in their publication. Only 16/46 (34.8%) of respondents reported they would be happy to use assay-level PS3/BS3 scores as provided by authors whilst 30/46 (65.2%) stated they would require independent review/repeating of the Brnich-style MAVE validation by another body (Figure 2A). When asked from which bodies they would accept such review, all 30/30 confirmed CanVIG-UK/CStAG would be acceptable, 25/30 (83.3%) would accept review by a relevant VCEP, and 10/30 (33.3%) would accept a review performed within their local GLH (Table S1).

**Figure 2:**
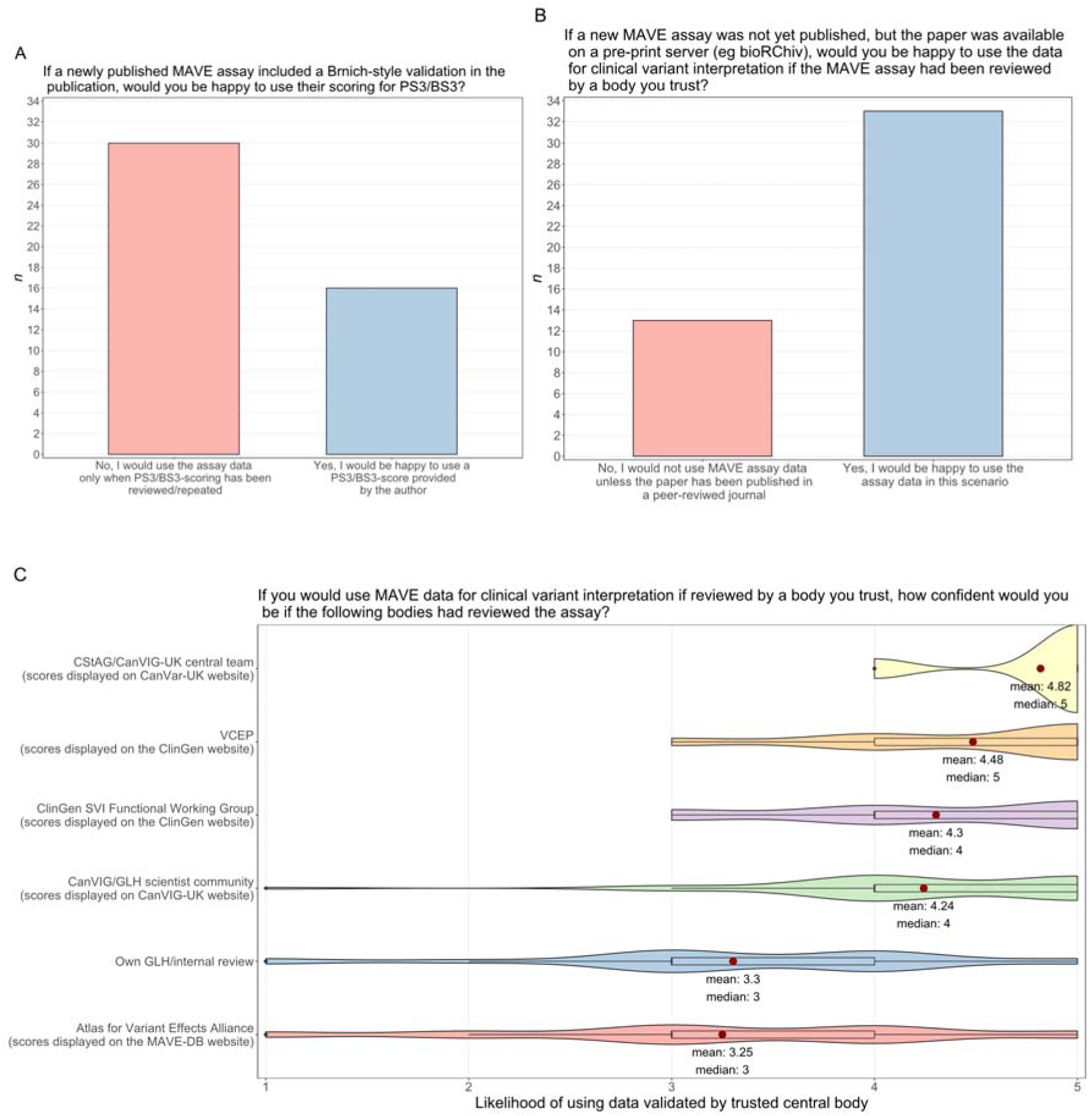
Results from survey questions on centralised clinical validation. *n* = Number of participants. A: Opinions on using assay data where validation performed only by the MAVE assay authors (n=46). B: Opinions on using pre-print assay data validated by a ‘trusted central body’ (n=46). C: Confidence of participants who would use validated pre-print assay data (n=33) which has been validated by each of the listed central bodies, scored from 1-5 (1=very unconfident, 5=very confident). Mean average and median scores are displayed; mean scores are represented by a red point for each group. One participant did not provide a score for the Atlas of Variant Effect Alliance group (n=32).

Participants were then asked if they would use unpublished MAVE data available on a pre-print archive such as bioRχiv (https://www.biorxiv.org/) provided that a ‘reputable body’ had reviewed and performed the Brnich-style validation (with display of the resultant assay-level PS3/BS3 scores on a corresponding website/portal). 33/46 (71.7%) confirmed readiness to use the MAVE data for clinical variant classification in this scenario (Figure 2B). Of these 33 participants, reported confidence (scored 0-5) was highest if the Brnich-style MAVE validation had been performed (i) centrally by the CanVIG-UK team/CStAG (median=5, mean=4.82) or via the collective CanVIG-UK clinical scientist community (median=4, mean=4.24), with assay-level PS3/BS3 scores being displayed on the CanVar-UK web platform in either instance (ii) the relevant VCEP (median=5, mean=4.48) or the ClinGen SVI Functional Working Group (median=4, mean=4.30) with assay-level PS3/BS3 scores being displayed on the ClinGen website in either instance (Figure 2C). The confidence in using the data was lower for Brnich-style MAVE validation performed by the AVE Alliance with assay-level PS3/BS3 scores being displayed on MaveDB (median=3, mean=3.25). Confidence was also lower for the Brnich-style MAVE validation being performed locally (median=3, mean=3.3).

### Gene and variant level data required additional to assay-level PS3/BS3 scores for clinical application of MAVE data

Finally, participants were asked whether, if provided with assay-level PS3/BS3 scores from a Brnich-style MAVE validation performed by a ‘trusted central body’, there were any gene-level or variant-level information that might influence their application of the prescribed assay-level PS3/BS3 scores for a given variant under evaluation. Scores (1 (low) to 5 (high)) were provided for the likelihood of influencing prescribed assay-level PS3/BS3 scores for two types of assay-level information, namely (i) type of cell line or (ii) type of assay, and for three types of variant-level information, namely (iii) number and (iv) consistency of replicate experiments, and (v) how close the absolute functional score was to the lower cut-off threshold. Overall, the majority of participants reported a low likelihood that either gene-level or variant-level factors would influence the assay-level PS3/BS3 scores assigned on Brnich-style MAVE validation when performed by a ‘trusted central body’ (Figure 3).

**Figure s3:**
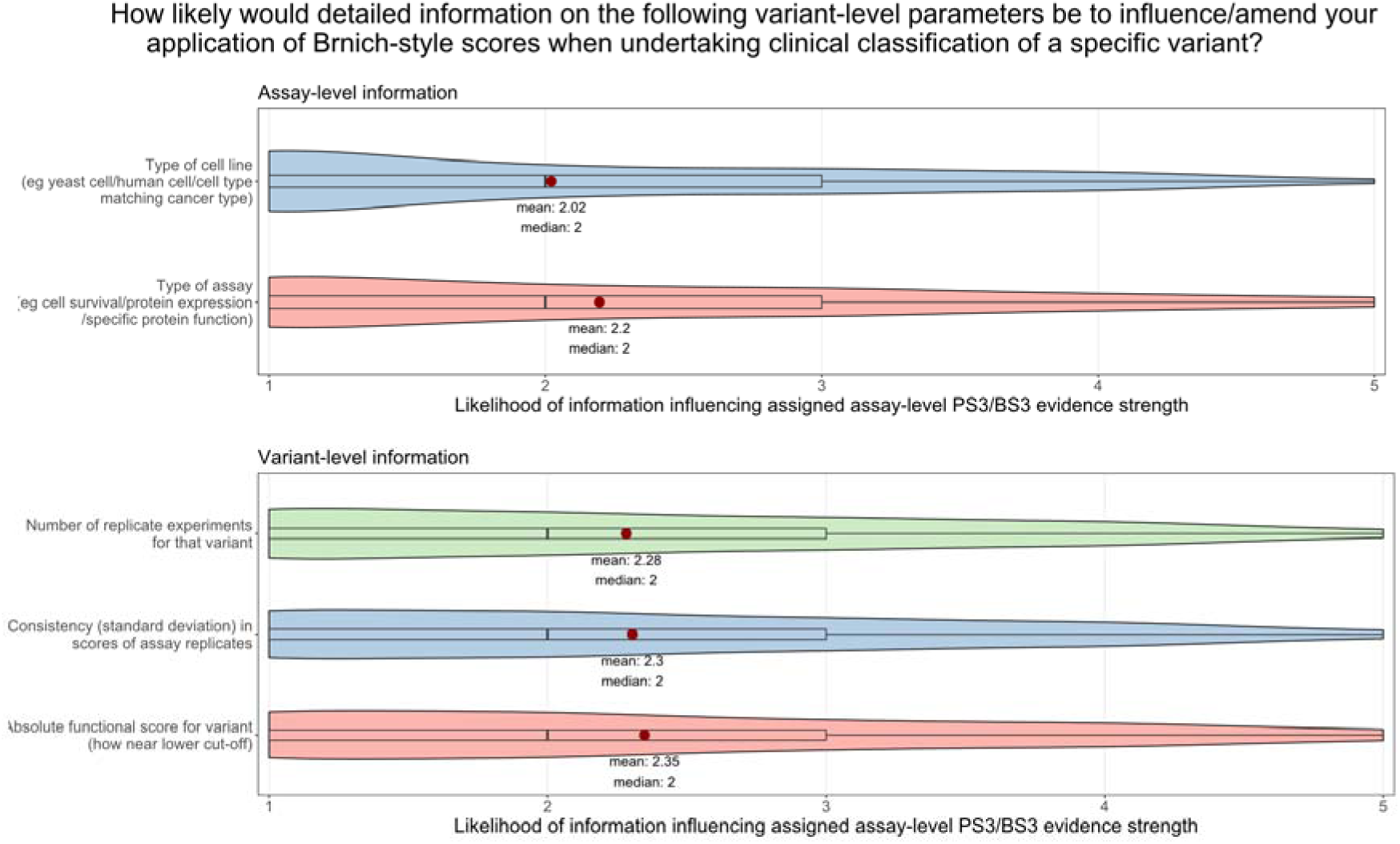
Participant scoring of the likelihood that assay-level information (type of cell line or type of assay) and/or variant-level information (number and consistency of replicates, and absolute functional score) will influence the PS3/BS3 evidence strength applied, scored from 1-5 (n=46, 1=low likelihood, 5=high likelihood). Mean average and median scores are displayed; mean scores are represented by a red point for each group.

## Discussion

We present survey data for 46 participants who regularly perform ACMG/AMP-based variant classification for clinical diagnostics, comprising NHS clinical diagnostic scientists (43/46 respondents) and clinical geneticists (3/46). The survey was initiated during a live meeting, preceded by verbal question-by-question presentation of the full survey. To optimise information, attention and comprehension by participants, we preceded delivery of the survey with presentations on (i) Brnich-style validation of MAVEs and (ii) available online repositories of MAVE data and (iii) description by authors of two recently published MAVEs. Responses from this relatively homogenous, NHS-based participant group, all regularly undertaking ACMG/AMP-based variant classification, offer useful insights regarding perspectives on and barriers to clinical application of MAVE data in a resource-constrained, real-world, clinical diagnostic setting.

Whilst almost half of participants had previously attempted a Brnich-style validation of a functional assay, only about 20% would proceed in attempting such a validation on a newly-published MAVE (specified as being of relevance to a gene they report upon). More than half of participants cited by way of explanation a lack of confidence regarding the Brnich-style validation methodology and uncertainty in assembling appropriate variant truthsets. The majority of participants also reported they lacked the necessary bioinformatics skills or resources for performing the analyses (for example for extracting truthset variants from ClinVar and lining up with assay results). Approximately two-thirds of participants cited time pressures as a barrier and one-third felt that it was beyond their remit. A number of free-text comments reflected the interrelated concerns regarding the Brnich-style validation being performed correctly and the impact of such validation on clinical-scientist time: “Clinical scientists are stretched too far already”, “It’s not the most intuitive calculation and would be very time-consuming to do”, “We need extra funding for personnel to perform this task”, “We haven’t defined a formal process to undertake this activity, we would do a superficial review using the Brinch flow chart [sic] rather than a specific assessment” and one unequivocal response of having “no idea where to start”.

Correlating with reluctance for local Brnich-style MAVE validations was a strong enthusiasm for a ‘trusted central body’ to take on this role (and make the relevant data available and accessible). Some manner of authorisation by such a ‘trusted central body’ would be deemed necessary by two-thirds of participants even for a MAVE for which the authors had themselves provided a Brnich-style validation and assay-level PS3/BS3 scoring in their publication. For the majority (33/46, 72%) of participants, authorisation by the ‘trusted central body’ superseded requirement for formal peer-reviewed publication (for example, data only submitted to a pre-publication archive).

There was strong endorsement for both the VCEPs and the ClinGen SVI Functional Working Group as potential ‘trusted central bodies’ who might provide Brnich-style validation of new MAVEs (under a hypothetical model that MAVE data and assay-level PS3/BS3 scores would then be made available on ClinVar). The lower enthusiasm for AVE Alliance as a ‘trusted central body’ may in part reflect less familiarity with the group, but also may reflect perceptions that MaveDB is less user-friendly (as currently configured) for a clinical diagnostic user. Evident from responses was strong support for extant national UK structures already operational in this realm. Namely the CanVIG-UK group (with its central team and oversight group CStAG), who already host a central online repository of Brnich-style MAVE validations performed by the CanVIG-UK central team and the collective CanVIG-UK clinical scientist community. By extension, participants also support data presentation on CanVar-UK, also hosted by CanVIG-UK and which provides variant-level MAVE data alongside other resources relevant to ACMG/AMP-based variant classification.

Under a hypothetical model of assay-level PS3/BS3 scores from Brnich-style validation being made available for all new MAVEs by some manner of ‘trusted central body’, it was then important to elicit which additional assay-level or variant-level annotations might be required alongside assay-level PS3/BS3 scores to support use of the MAVE data for ACMG/AMP-based variant classifications. The majority of participants reported there was a low likelihood that further review of assay-level information would cause them to alter the assay-level PS3/BS3 scores provided by the ‘trusted central body’, as reflected by one of the free-text comments: “If validated by a recognised trusted group we would not amend the weighting the functional scores determined by that group”. However, caution was voiced by some participants around assay-level issues, for example the need for a guarantee that the assay “includes ALL mechanisms of disease e.g. missense protein function and potential for LOF via abnormal splicing”. There was a modestly higher mean likelihood that variant-level data annotations may influence assay-level PS3/BS3 scoring, in particular where the variant’s assay score lay close to the absolute assay threshold. One participant cautioned they “would definitely be more cautious for variants at start / end of exons”, whilst another emphasised that they would “review publicly available experimental parameters especially if the functional scores applicable according to Brinich seems at odds with other lines of evidence on the specific variant”. It would appear overall that whilst participants would largely rely on the assay-level PS3/BS3 scores provided by the ‘trusted central body’, they would value variant-level data on absolute scores and replicates being provided alongside (to enable review for equivocal variants or particular scenarios). This mirrors findings of polls of other stakeholder groups regarding clinical application of MAVE data, namely concurrent appetite for wanting just the validated assay-level PS3/BS3 score, but also wanting access to extensive variant-level data to accompany the score^18,19^.

### Limitations

Those attending a CanVIG-UK meeting focused on MAVEs may be a subgroup of CanVIG-UK members skewed by interest in and familiarity with MAVEs. Thus, participants who responded to the poll may be more likely to be actively engaged with using functional evidence and the issues surrounding clinical validation of MAVE data. Only approximately half of meeting attendees submitted a response, with non-responders likely including clinical geneticists and trainees who do not regularly perform ACMG/AMP-based variant classification but also potentially reflecting individuals less interested, willing or available to participate in the poll. By virtue of their presence at a CanVIG-UK meeting, participants are also likely to have a positive attitude towards CanVIG-UK as an organisation and its current/potential roles for MAVE validation.

It is possible that participants less familiar with functional assays may have misunderstood or lacked knowledge regarding some questions (for example, relating to Brnich-style methodology or the various central bodies), albeit that we had sought to mitigate this with the pre-survey introductory talks and verbal presentation of the survey (and restriction to participants who regularly undertake ACMG/AMP-based variant classification).

The survey findings benefit from the relative homogeneity of participants but necessarily reflects the specific context of UK NHS clinical scientists and clinical geneticists working in cancer susceptibility genetics. A number of the responses relate to UK-specific resources provided by and activities undertaken by the CanVIG-UK group (with its central team and oversight group CStAG). No groups equivalent to CanVIG-UK exist for other UK disease subspecialities, so responses might differ regarding validation of MAVEs if performing the survey within the UK rare disease or cardiac genetics communities.

## Conclusions and recommendations

Our survey provides compelling evidence that (lack of) availability of ‘trusted’ Brnich-style clinical validations represents a significant potential barrier to clinical application of new MAVE data. The majority of clinical scientists report they would be unwilling to use clinical validation metrics provided by MAVE authors, but also report that they lack the capacity, confidence and bioinformatics resources for undertaking the validation locally in their laboratories. This survey relates to the current methodology for Brnich-style MAVE validation (generating assay-level PS3/BS3 scores): it is likely that any new methodologies also incorporating variant-specific validation metrics and/or for combining multiple MAVEs for a given gene will prove even more challenging for local laboratories and/or non-experts to perform.

From our survey findings, we identified the following recurring themes which form a set of recommendations from the collective CanVIG-UK clinical scientist community regarding the integration and use of MAVE data within variant classification:

1. **A trusted central body/bodies should take responsibility for Brnich-style MAVE validation (rather than individualised laboratories), on account of (i) consistency/robustness of approach and (ii) local manpower implications.**

The survey reflects broad support for a number of organisations who might take on this role of a ‘trusted central body’: VCEPs, the ClinGen SVI Functional WG, or the AVE Alliance. However the ClinGen SVI Functional WG and AVE Alliance, whilst developing and advising on validation methodology, have not to date undertaken Brnich-style validations of individual MAVEs. Whilst VCEPs will variously sanction application of data from specified assays at specified evidence scores, the rationale, validation methodology and truthsets used by the VCEP are seldom explicitly provided and are likely widely variable. Furthermore, VCEPs are volunteer-led and operate in delivery cycles of per-gene ACMG/AMP framework specifications which must be centrally approved before release; accordingly, they are currently not typically well-placed to deliver timely, responsive Brnich-style validations of new MAVEs. The survey reflects approval and trust for a ‘mid-scale’ model of a national trusted central body, as exemplified by CanVIG-UK, resourced by a small central team which can then leverage and coordinate input from the collective clinical diagnostic community.

2. **Clinically-facing displays of MAVE data should include the absolute assay score (variant-level) and the assay-level evidence strength resulting from centralised Brnich-style MAVE validation.**

Platforms intending to provide clinically-facing displays of MAVE data should be readily accessible and offer user-friendly, variant-level look-up. Incorporation onto platforms already used for querying other data elements relating to ACMG/AMP-based variant classification (such as ClinVar or CanVar-UK) is likely to provide the preferred workflow for accessing MAVE data, rather than requiring clinical professionals to access an additional, distinct resource.

3. **Assay-level and variant-level information should be made available for all variants included in the MAVE publications, alongside or readily linked to the clinically-facing displays of MAVE data.**

Although deemed by the majority of respondents unlikely to influence clinical application of assay-level PS3/BS3 scores as provided by a central, “trusted” body, it appeared overall that routine availability of these data to clinical users would be of value to support contextualisation of MAVE results in equivocal or unusual scenarios.

## Acknowledgments

We would like to thank the CanVIG-UK members who participated in this survey. A full list of all CanVIG-UK consortium members and their affiliations appears in the Supplemental Material. SA and CFR are supported by CG-MAVE, CRUK Programme Award [EDDPGM-Nov22/100004]. AG and HH are supported by CRUK Catalyst Award CanGene-CanVar [C61296/A27223].

## Author Contributions

Conceptualization: CT, DJA, GMF. Funding acquisition: CT, DJA, GMF. Methodology: CT, AG, CFR, MD, GJB, RR, AC, JF, BF, SPS, JG, JP, TMcD, KS, HH, TMcV, RMV, ABS. Investigation: CT, AG, SA. Data curation: SA. Formal analysis: SA. Visualisation: SA. Writing – original draft: CT, SA. Writing – review and editing: All authors.

## Data availability

The published article includes all datasets generated or analysed during this study.

