## Supplementary Material for "Validating data from Multiplex Assays of Variant Effect (MAVEs): A CanVIG-UK National Survey of NHS Clinical Scientists"

### Supplemental Methods

Question content and electronic format/usability was iterated through several expert groups knowledgeable in clinical variant interpretation, to ensure questions were clear for participants with a clinical background. These groups included:

- The CanVIG-UK Steering and Advisory Group, comprising 10 senior clinical scientists and 3 consultant clinical geneticists, providing representation from across all English Genomic Laboratory Hubs, Scotland, Wales, and Ireland.
- Members of the AVE-Alliance Clinical Variant Interpretation Workstream (https://www.varianteffect.org/workstreams), who have experience in running similar surveys regarding use and validation of MAVE data.[1]

Participants were self-selected from attendees at the CanVIG-UK monthly meeting on 15th November 2024. These are closed meetings attended by CanVIG-UK members. The participant group was further selected based on self-declared involvement of the participant in variant ACMG/AMP classification of variants. Those who responded 'yes' to this question were included in the final results.

The survey link, prepared using SurveyMonkey (http://www.surveymonkey.com), was circulated to attendees both during the meeting and afterwards by email, and remained live until 20th November 2024. To provide all participants with equivalent base-level of knowledge regarding MAVE data, the meeting consisted of several short talks to introduce the topics surveyed including validation of MAVE data and the Brnich et al. methodology[2], and existing MAVE data resources such as MaveDB, ClinVar, and the Atlas of Variant Effect (AVE-Alliance)[3-5]. The investigators also provided a demonstration of the survey questions and possible responses live during the meeting and fielded questions from participants to mitigate the possibility of participants misinterpreting the questions while responding.

Participation in the survey was voluntary. No incentives were offered for completion of the survey. Prior to the survey being circulated in the meeting, and in the subsequent email circulation with the meeting minutes, participants were informed of the length of time needed to complete the survey (around 5 minutes), and that their data would be stored by the investigators (Prof. Clare Turnbull and the CanVIG-UK central team at the Institute of Cancer Research) for review and analysis. Participants were provided with information regarding the purpose of the study and how their responses would be used, which was stated to be in learning how functional data is being used across Genomic Laboratory Hubs (GLHs) and determining which data from these studies would be useful for clinical presentation of MAVE data.

The survey comprised 11 questions, plus a free-text comment box, across the topics of current validation methods by diagnostic laboratories for MAVE data, centralised vs local review of MAVE data, and types of assay-level and variant-level information which may be considered when applying MAVE data as part of variant classification. Questions were not randomised, and were presented on a single scrollable page. 8/11 questions were mandatory, and required selection of at least one option. 3/11 questions were optional, as these were reliant on specific responses to previous questions (such as a previous 'Yes' response). All questions included either a not applicable option or an 'Other' option, with free-text comment box provided at the end of the survey. Respondents were able to save their progress and return to the survey to change answers or complete the survey at any point before the survey closed. Completion was reviewed after the questionnaires were submitted.

Personal identifiable information was not collected as part of this survey. IP address was collected for the purpose of identifying unique visitors or duplicate submissions, and this information was not stored after confirming no duplicate submissions were made. A time period of 24 hours was instated during which time a second response from the same IP address would be deemed to be duplicate. If this was the case, the most recent response would have be retained, and the previous response removed. Survey responses and date of survey response were downloaded from SurveyMonkey and held for the duration of study analysis on a secure drive with access provided only to the investigators.

The survey received a total of 48 responses of the 86 attendees present at the CanVIG-UK meeting (recruitment rate: 55.8%), all of which were from unique IP addresses. All 48/48 responses were complete. 46/48 responses were from individuals who confirmed regular undertaking of ACMG/AMP classification of variants, and these 46 results were retained for further analysis.

#### Supplemental Note 1: Full CanVIG-UK Membership and Affiliations

C. Turnbull[1,49], A. Garrett[1,47], L. Loong[1], S. Choi[1], B. Torr[1], S. Allen[1], M. Durkie[2], A. Callaway[3], J. Drummond[4], G.J. Burghel[5,86], R. Robinson[6], I.R. Berry[65], A.J. Wallace[5], D.M. Eccles[7, 8], M. Tischkowitz[13], S. Ellard[9], H. Hanson[1,88,89], E. Baple[10,11], D.G. Evans[5,30], E. Woodward[5,30], F. Lalloo[5,30], S. Samant[33], A. Lucassen[57,14,15], A. Znaczko[44], A. Shaw[23], A. Ansari[34], A. Kumar[21], A. Donaldson[53], A. Murray[19], A. Ross[18], A. Taylor-Beadling[22], A. Taylor[18], A. Innes[25], A. Brady[29], A. Kulkarni[23], A.C. Hogg[5,] A. Ramsay Bowden[18], A. Hadonou[47], B. Coad[16], B. McIldowie[19], B. Speight[18], B. DeSouza[47], B. Mullaney[3], C. McKenna[62], C. Brewer[44], C. Olimpio[18], C. Clabby[40], C. Crosby[47], C. Jenkins[42], C. Armstrong[33], C. Bowles[9], C. Brooks[22], C. Byrne[62], C. Maurer[4], D. Baralle[57], D. Chubb[1], D. Stobo[34], D. Moore[35], D.O'Sullivan[33], D. Donnelly[62], D. Randhawa[24], D. Halliday[41] , E. Atkinson[50], E. Rauter[24], E. Johnston[38], E. Maher[8], E. Sofianopoulou[17], E. Petrides[23], F. McRonald[43], F. Pelz[51], I. Frayling[83,87], G. Corbett[62], G. Rea[62], H. Clouston[5], H. Powell[31], H. Williamson[52], H. Carley[47], H.J.W. Thomas[26], I. Tomlinson[63], J. Cook[46], J. Tellez[32], J. Whitworth[18], J. Williams[49], J. Murray[35], J. Campbell[27], J. Tolmie[33], J. Field[38], J. Mason[64], J. Burn[31], J. Bruty[18], J. Callaway[3], J. Grant[34], J. Del Rey Jimenez[47], J. Pagan[35], J. VanCampen[24], J. Barwell[53], K. Monahan[29], K. Tatton-Brown[16,] K.R. Ong[63], K. Murphy[33], K. Andrews[18,] K. Mokretar[23], K. Cadoo[48], K. Smith[52], K. Baker[8], K. Brown[24], K. Reay[64], K. McKay Bounford[34], K. Bradshaw[38], K. Russell[65], K. Stone[23], K. Snape[16], L. Crookes[5], L. Reed[21], L. Yarram-Smith[65], L. Cobbold[47], L. Walker[39], L. Walker[41], L. Hawkes[16], L. Busby[22], L. Izatt[23], L. Kiely[22,] L. Hughes[64], L. Side[56], L. Sarkies[18], K.-L. Greenhalgh[28], M. Shanmugasundaram[63], M. Duff[40], M. Bartlett[29], M. Watson[3], M. Owens[9], M. Bradford[54], M. Huxley[64], M. Slean[33], M. Ryten[23], M. Smith[55], M. Ahmed[21], N. Roberts[2], O. Middleton[33], P. Tarpey[4], P. Logan[62], P. Dean[3], P. May[24], P. Brace[21], R. Tredwell[38], R. Harrison[37], R. Hart[63], R. Martin[31], R. Nyanhete[3], R. Wright[2], R. Martin[62], R. Davidson[34], R. Cleaver[45], S. Talukdar[16], S. Butler[64], J. Sampson[19], S. Ribeiro[49], S. Dell[46], S. Mackenzie[32], S. Hegarty[62], S. Albaba[5], S. McKee[36], S. Palmer-Smith[19], S. Heggarty[62], S. MacParland[62], S. Greville-Heygate[49], S. Daniels[4],S. Prapa[18],S. Abbs[4], S. Tennant[33], S. Hardy[43], S. MacMahon[49], T. McVeigh[49], T. Foo[49], T. Bedenham[42], T. Cranston[42,] T. McDevitt[40], V. Clowes[29], V. Tripathi[23], V. McConnell[62], N. Woodwaer[45], Y. Wallis[64], Z. Kemp[49], G. Mullan[62], L. Pierson[62], L. Rainey[62], C. Joyce[59], A. Timbs[41], A-M. Reuther[3], B. Frugtniet[47], B. DeSouza[25], C. Husher[3], C. Lawn[22], C. Corbett[63], D. Nocera-Jijon[16], D. Reay[31], E. Cross[3], F. Ryan[3], H. Lindsay[6], J. Oliver[6], J. Dring[63], J. Spiers[65], J. Harper[23], K. Ciucias[34], L. Connolly[60], M. Tsang[62], R. Brown[6], S. Shepherd[32], S. Begum[16], S. Daniels[3], T. Tadiso[16], T. Linton-Willoughby[4], H. Heppell[35], K. Sahan[61], L. Worrillow[6], Z. Allen[22], C. Watt[34],M. Hegarty[62], R. Mitchell[6], R. Coles[66], G. Nickless[23], E. Cojocaru[49], I. Doal[64], F. Sava[64], C. McCarthy[62], R. Jeeneea[63], D. Goudie[20], M. McConachie[20], S. Botosneanu[5], G. Kavanaugh[1], K. Russell[10], C. Sherlaw[63], O. Tsoulaki[46], C. Forde[5], E. Petley[63], A-B. Jones[1], K. Oprych[16], S. Pryde[67], Z. Hyder[5], N. Elkhateeb[18], R. Braham[21], L. Hanington[41], C. Huntley[1], R. Irving[51], A. Sadan[23], M. Ramos[22], C. Elliot[35], D. Wren[22], D.Lobo[35], J. McLean[68], D. May[18], L. Kearney[48], T. Campbell[38], K. Asakura[68], L. Alwadi[19], R. O'Shea[48], J. Gabriel[42], L. Chiecchio[3], P. Bowman[44], L.A. Sutton[48], C. Walsh[23], V. Cloke[69], D. Ucanok[37], J. Davies[65], B. Pleasance[65], E. Maguire[6], A. Whaite[70], S. Best[71], S. Westbury[72], A. Logan[62], D. Navarajasegaran[71], A. Bench[35], P. Wightman[34], A. Cartwright[2], E. Higgs[41], J.Bott[42], H. Whitehouse[5], J. Stevens[58], D. Martin[37], J. Dunlop[68], S. Thomas[73], C. Sau[68], S. Farndon[74], N. Coleman[48], P. Angelini[49], M. Duff[59], H. Massey[35], C. Rowlands[1], C. Garcia-Petit[68], K. Gillespie[68], A. Alder[68], E. Middleton[68], C. Cassidy[75], N. Orfali[48], A. Webb[3], A. Luharia[64], N. Walker[33], J. Charlton[71], A. Andreou[47], J. Peddie[68], M. Khan[23], L. Wilkinson[31], H. Bezuidenhout[47], M. Edis[4], A. Callard[29], P. Ostrowski[76], P. Moverley[51], K. Bean[73], A. Dunne[48], A. Moleirinho[23], S. Waller[5], K. Cox[47], L. Greensmith[28], A. Brittle[5], N. Gossan[5], L. Freestone[4], C. Shak[77], T. Langford[75], Y. Clinch[21], H. Livesey[51], S. Borland[46], A. Joshi[47], K. Wall[77], A. Whitworth[46], A.Wilsdon[37], K. Edgerley[72], S. Pugh[5], N. Chrysochoidi[3], S. Mutch[38], C. McMullan[5], Y. Johnston[78], M. Muraru[77], A. May[77], R. Begum[77], C. Smith[44], R. Patel[47], I. Bhatnagar[79], A. Taylor[62], D. Brown[69], J. Willan[46], S. Taylor[48], K. Jones[21], K. Cox[21], C. Ramsden[75], O. Taiwo[49], J. Jaudzemaite[47], R. Sharmin[47], L. Young[34], C.O'Dubhshlaine[35], L. McSorley[80], S. Lillis[23], P. Alexopoulos[23], E. Mortensson[65], L. Kingham[4], R. Moore[18], M. Kosicka-Slawinska[81], S. Aslam[65], R. Wells[82], A. Carter[82], H. Warren[6], E. Rolf[49], H. Reed[75], L. Pearce[19], D. Lock[6], F. Ali[66], A. Kolozi[84], N. White[48], D. Wood[34], C. Hayden[25], W. Cheah[58], J. Sims[6], R. Heron[46], J. Sibbring[4], L. Elmhirst[35], L. Mavrogiannis[6], K. Oakhill[4], L. Wang[28], A. Singh[47], K. Doal[21], L. Kettle[63], R. Salmon[63], G. Thodi[48], C. O'Brien[48], C. Wragg[85], N. Mannion[34], S. Chu[63], M. Ukash[75], V. Steventon-Jones[46], J. Fairley[84], H. Northen[18], D. Babu[75], L. Donaghy[34], J. Jimmy[48], B. Matharu[18], J. Beasley[42], S. Waller[75], C. Batterton[63], G. Baker[4], J. Trotman[4], L. Jackson[11], A. Visavadia[63], M. Domeradzka[49], M. Slater[22], K. Annesley[18], C. Andrews[1], J. Doughty[18], E. Wall[63], S. Morosini[46], E. Hanney[60], H. Cheema[46], H. Skinner[46], A. Western[46], M. Cabes[41], J. Grant[41], N. Brodaczewska[25], L. Gilroy[35], E. Phillips[4], R. Lane[41], E. Higgs[21], G, Afifi[18], N. Ali[36], M. Gordan[62], L. Clarke[77], R. Aungraheeta[65], L. Redford[22], I. Richards[63], R. Price[63], C. Quinn[62], G. Beard[24]

[1] Division of Genetics and Epidemiology, Institute of Cancer Research, Sutton, UK

[2] Sheffield Diagnostic Genetics Service, NEY Genomic Laboratory Hub, Sheffield Children's NHS Foundation Trust, Sheffield, UK

[3] Wessex Genetics Laboratory Service, University Hospital Southampton NHS Foundation Trust, Salisbury, UK

[4] East Genomic Laboratory Hub, Cambridge University Hospitals Genomic Laboratory, Cambridge University NHS Foundation Trust, Cambridge, UK

[5] Manchester Centre for Genomic Medicine and NW Laboratory Genetics Hub, Manchester University Hospitals NHS Foundation Trust, Manchester, UK

[6] The Leeds Genetics Laboratory, NEY Genomic Laboratory Hub, Leeds Teaching Hospitals NHS Trust, Leeds, UK

[7] Cancer Sciences, Faculty of Medicine, University of Southampton, Southampton, UK

[8] Human Genetics and Genomic Medicine, Faculty of Medicine, University of Southampton, Southampton, UK

[9] Department of Molecular Genetics, Royal Devon and Exeter NHS Foundation Trust, Exeter, UK

[10] Genomics England, London, UK

[11] University of Exeter Medical School, Exeter, UK

[12] Division of Evolution & Genomic Sciences, The University of Manchester

[13] Department of Medical Genetics, National Institute for Health, Research Cambridge Biomedical Research Centre, University of Cambridge, Cambridge, UK

[14] Wessex Clinical Genetics Service, University Hospital Southampton NHS Foundation Trust, Southampton, UK

[15] Clinical Ethics and Law Unit, University of Southampton, Southampton, UK

[16] Department of Clinical Genetics, St. George's University Hospitals NHS Foundation Trust, London, UK

[17] Public Health and Primary Care, Clinical Medicine, University of Cambridge, Cambridge, UK

[18] Cambridge University Hospitals NHS Foundation Trust, Cambridge, UK

[19] All Wales Medical Genomics Service, Wales Genomics Health Centre, Cardiff UK

[20] East of Scotland Regional Genetics Service, Level 6, Ninewells Hospital, Dundee

[21] Great Ormond Street Hospital for Children NHS Foundation Trust, London, UK

[22] North Thames Genomic Laboratory Hub, Great Ormond Street Hospital for Children NHS Foundation Trust, London, UK

[23] Department of Clinical Genetics, Guy’s and St Thomas’ NHS Foundation Trust, London, UK

[24] South East Genomic Laboratory Hub, Guy’s and St Thomas’ NHS Foundation Trust, London, UK

[25] Genomic Medicine Service, Imperial College Healthcare NHS Trust, London, UK

[26] Faculty of Medicine, Department of Surgery & Cancer, Imperial College London, London, UK

[27] Institute of Neurology, UCL Queen Square Institute of Neurology, London, UK

[28] Liverpool Women’s NHS Foundation Trust, Liverpool, UK

[29] London North West University Healthcare NHS Trust, London, UK

[30] Division of Evolution and Genomic Sciences, School of Biological Sciences, Faculty of Biology Medicine and Health, The University of Manchester, Manchester, UK.

[31] The Newcastle upon Tyne Hospitals NHS Foundation Trust, Newcastle upon Tyne, UK

[32] North East and Yorkshire Genomic Laboratory Hub, The Newcastle upon Tyne Hospitals NHS Foundation Trust, Newcastle upon Tyne, UK

[33] NHS Grampian, Aberdeen, UK

[34] NHS Greater Glasgow and Clyde, Glasgow, UK

[35] NHS Lothian, Edinburgh, UK

[36] Northern Ireland Regional Genetics Service, Belfast Health & Social Care Trust, Belfast, UK

[37] Nottingham University Hospitals NHS Trust, Nottingham, UK

[38] East Midlands and East of England Genomics Laboratory, Nottingham University Hospitals NHS Trust, Nottingham, UK

[39] University of Otago, Otago, New Zealand

[40] Our Lady's Children's Hospital, Crumlin, Dublin, Ireland

[41] Clinical Genetics, Oxford University Hospitals NHS Foundation Trust, Oxford, UK

[42] West Midlands, Oxford and Wessex Genomic Laboratory Hub, Oxford University Hospitals NHS Foundation Trust, Oxford, UK

[43] Public Health England, London, UK

[44] Royal Devon and Exeter NHS Foundation Trust, Exeter, UK

[45] Royal Free London NHS Foundation Trust, London, UK

[46] Sheffield Children's NHS Foundation Trust, Sheffield, UK

[47] St George’s University Hospitals NHS Foundation Trust, London, UK

[48] St James’s Hospital, Dublin, Ireland

[49] Cancer Genetics Unit, The Royal Marsden NHS Foundation Trust, Sutton, London, UK

[50] Trinity College Dublin, The University of Dublin, Ireland

[51] University Hospital of Wales, Cardiff and Vale University Health Board, Cardiff, UK

[52] University Hospitals Bristol NHS Foundation Trust, Bristol, UK

[53] University Hospitals of Leicester NHS Trust, Leicester, UK

[54] University Hospitals of Plymouth NHS Trust, Plymouth, UK

[55] University of Manchester, Manchester, UK

[56] Wessex Clinical Genetics Service, Princess Anne Hospital, Southampton, UK

[57] Faculty of Medicine, University of Southampton, Southampton, UK

[58] University Hospital Southampton NHS Foundation Trust, Southampton, UK

[59] Cork University Hospital, Cork, Ireland

[60] Children’s Health Ireland (CHI), Crumlin, Dublin, Ireland

[61] The Ethox Centre, Oxford, UK

[62] Belfast Health & Social Care Trust, Belfast, UK

[63] Birmingham Women's and Children's NHS Foundation Trust, Birmingham, UK

[64] Central and South Genomic Laboratory Hub, Birmingham Women's and Children's NHS Foundation Trust, Birmingham, UK

[65] Bristol Genetics Laboratory, Pathology Sciences, Southmead Hospital, North Bristol NHS Trust, Bristol, United Kingdom

[66] Northwick Park Hospital, Watford Rd, Harrow, UK

[67] Chapel Allerton Hospital, Chapeltown Rd, Leeds, UK

[68] NHS Tayside, UK

[69] South East Scotland Genetic Service, Western General Hospital, Edinburgh, UK

[70] Liverpool Centre for Genomic Medicine, Liverpool Women's NHS Foundation Trust, Liverpool, UK

[71] King's College Hospital, London, UK

[72] University Hospital Bristol and Weston NHS Foundation Trust, Bristol, UK

[73] Sheffield Teaching Hospital NHS Foundation Trust, Sheffield, UK

[74] Bristol Royal Hospital for Children, Bristol. UK

[75] Manchester University Foundation Trust, Manchester, UK

[76] North East Thames Regional Genetics Service, London, UK

[77] West Midlands Regional Genetics Laboratory, Birmingham Women's Hospital, Birmingham, UK

[78]West of Scotland Centre for Genomic Medicine, Queen Elizabeth University Hospital, Glasgow, UK

[79]Oxford Centre for Genomic Medicine, Oxford University Hospitals NHS Foundation Trust, Oxford, UK

[80]St Vincent's Hospital Group, Elm Park, Dublin, Ireland

[81]North West Thames Regional Genetics Service, St. Mark's Hospital, Harrow, UK

[82]Royal Liverpool University Hospital Trust, Liverpool, UK

[83]Inherited Tumour Syndromes Research Group, Division of Cancer and Genetics, School of Medicine, Cardiff University

[84]GenQA

[85]South West Genomic Laboratory Hub, Bristol Genetics Laboratory, Southmead Hospital, Bristol, UK

[86]Division of Cancer Sciences, School of Medical Sciences, Faculty of Biology, Medicine and Health, The University of Manchester

[87]St Mark's Centre for Familial Intestinal Cancer, St Mark's Hospital, Central Middlesex, Acton Lane, Park Royal

[88]Peninsula Regional Genetics Service, Royal Devon University Healthcare NHS Foundation Trust, Exeter, UK

[89]Department of Clinical and Biomedical Sciences, University of Exeter Medical School, Exeter, United Kingdom

**Table S1: All survey questions and full results**

| Question | Options | | Number of responses |
|---|---|---|---|
| 1 What is your current role? | Pre-registration clinical scientist | | 2 |
| | Clinical Scientist | | 28 |
| | Principal clinical scientist | | 13 |
| | Clinical genetics registrar | | 0 |
| | Clinical genetics consultant | | 3 |
| | Genetic counsellor | | 0 |
| | Other | | 0 |
| 2 Do you undertake ACMG/AMP classification of variants regularly (more than once a month) within your role (even if not formally signing out lab reports)? | Yes | | 46 |
| | No | | 0 |
| 3 Have you ever undertaken a Brnich-style validation of a functional assay? | Yes | | 20 |
| | No | | 26 |
| 4 If there was a new MAVE published that was useful for a gene tested by your GLH that you report on (eg FH, VHL, PTEN), would you attempt a Brnich-style validation locally? | Yes, I would attempt myself | | 9 |
| | No, there is someone else in the GLH who undertakes Brnich-style validations for assays for germline cancer genes | | 2 |
| | No, our GLH would typically await validation by another body such as CanVIG/VCEP | | 28 |
| | No, I am unsure of local processes | | 7 |
| 5 If you answered No to Q4 about conducting a Brnich, which of the following are factors (select all that apply): | Not confident enough in Brnich methodology to use my results | | 21 |
| | Not confident in defining truthsets | | 17 |
| | Too time-consuming | | 24 |
| | Lack of bioinformatics support (e.g. for extracting truthset variants from ClinVar and lining up with assay results) | | 21 |
| | Beyond my remit | | 11 |
| | I would be happy doing it but it is the role of someone else in the GLH/lab | | 1 |
| | Other | | 4 |
| 6 If a newly published MAVE assay included a Brnich-style validation in the publication, would you be happy to use their scoring for PS3/BS3? | Yes, I would be happy to use a PS3/BS3-score provided by the author | | 16 |
| | No, I would use the assay data only when PS3/BS3-scoring has been reviewed/repeated | | 30 |
| 7 If you answered No to Q6, select each of the below options for which you would use the assay data if that body had reviewed/repeated PS3/BS3-scoring | Own GLH | | 10 |
| | CStAG/CanVIG-UK | | 30 |
| | VCEP | | 25 |
| 8 If a new MAVE assay was not yet published, but the paper was available on a pre-print server (eg bioRChiv), would you be happy to use the data for clinical variant interpretation if the MAVE assay had been reviewed by a body you trust? | Yes, I would be happy to use the assay data in this scenario | | 33 |
| | No, I would not use MAVE assay data unless the paper has been published in a peer-reviwed journal | | 13 |
| 9 If you answered Yes to Q8, how confident would you be in using such assay data if the following bodies had reviewed the assay (where 1 is very unconfident, and 5 is very confident) | CStAG/CanVIG-UK central team (scores displayed on CanVar-UK website) | 1 | 0 |
| | | 2 | 0 |
| | | 3 | 0 |
| | | 4 | 6 |
| | | 5 | 27 |
| | CanVIG/GLH scientist community (scores displayed on CanVIG-UK website) | 1 | 1 |
| | | 2 | 0 |
| | | 3 | 3 |
| | | 4 | 15 |
| | | 5 | 14 |
| | Own GLH/internal review | 1 | 3 |
| | | 2 | 2 |
| | | 3 | 13 |
| | | 4 | 12 |
| | | 5 | 3 |
| | ClinGen SVI Functional Working Group (scores displayed on the ClinGen website) | 1 | 0 |
| | | 2 | 0 |
| | | 3 | 6 |
| | | 4 | 11 |

| | | | | |
|---|---|---|---|---|
| | | | 5 | 16 |
| | | VCEP (scores displayed on the ClinGen website) | 1 | 0 |
| | | | 2 | 0 |
| | | | 3 | 4 |
| | | | 4 | 9 |
| | | | 5 | 20 |
| | | Atlas for Variant Effects Alliance (AVE Alliance) (scores displayed on the MAVE-DB website) | 1 | 3 |
| | | | 2 | 4 |
| | | | 3 | 11 |
| | | | 4 | 10 |
| | | | 5 | 4 |
| | | | Missing | 1 |
| 10 | If your Brnich-style scores (for example PS3_mod; BS3_strong) have been formally generated for a published assay by a body you trust, how likely would detailed information on the following assay-level parameters be to influence/amend your application of those Brnich-scores when undertaking clinical classification of a specific variant? (1=Low Likelihood, would go with the Brnich-style scoring for pathogenicity/benignity for the specified ranges of assay score; 5=High Likelihood, would individually amend the Brnich-style scoring for pathogenicity/benignity based on my own interpretations of these assay-level/variant-level factors) | Type of cell line (eg yeast cell/human cell/cell type matching cancer type) | 1 | 22 |
| | | | 2 | 9 |
| | | | 3 | 8 |
| | | | 4 | 6 |
| | | | 5 | 1 |
| | | Type of assay (eg cell survival/protein expression/specific protein function) | 1 | 19 |
| | | | 2 | 10 |
| | | | 3 | 9 |
| | | | 4 | 5 |
| | | | 5 | 3 |
| 11 | If your Brnich-style scores (for example PS3_mod; BS3_strong) have been formally generated for a published assay by a body you trust, how likely would detailed information on the following variant-level parameters be to influence/amend your application of those Brnich-scores when undertaking clinical classification of a specific variant? (1=Low Likelihood, would go with the Brnich-style scoring for pathogenicity/benignity for the specified ranges of assay score; 5=High Likelihood, would individually amend the Brnich-style scoring for pathogenicity/benignity based on my own interpretations of these assay-level/variant-level factors) | Absolute functional score for variant (how near lower cut-off) | 1 | 15 |
| | | | 2 | 13 |
| | | | 3 | 8 |
| | | | 4 | 7 |
| | | | 5 | 3 |
| | | Number of replicate experiments for that variant | 1 | 16 |
| | | | 2 | 12 |
| | | | 3 | 9 |
| | | | 4 | 7 |
| | | | 5 | 2 |
| | | Consistency (standard deviation) in scores of assay replicates | 1 | 15 |
| | | | 2 | 13 |
| | | | 3 | 9 |
| | | | 4 | 7 |
| | | | 5 | 2 |
